# Multi-dimensional Social Adversity and Mortality Outcome in People Living With HIV and Heart Failure: Insights from NYC Health + Hospitals HIV-Heart Failure cohort

**DOI:** 10.64898/2026.01.12.26343963

**Authors:** Yi-Yun Chen, Pawel Borkowski, Luca Biavati, Natalia Nazarenko, Matthew Parker, Amrin Kharawala, Coral Vargas-Pena, Ishmum Chowdhury, Joshua Bock, Vibhor Garg, Shivang Bhakta, Robert Faillace, Leonidas Palaiodimos, Wen-Chih Hank Wu, Elena Salmoirago-Blotcher, Sebhat Erqou, Chris T. Longenecker

## Abstract

**Background:** Heart failure (HF) is an increasingly common comorbidity among people living with HIV (PLHIV), complicating care and heightening vulnerability of this population to social adversity (SA). However, the impact of different SA domains on outcomes in this population remains poorly understood.

**Methods:** We analyzed data on PLHIV with HF from the NYC Health + Hospitals HIV-Heart Failure cohort. Baseline multidimensional SA was assessed by licensed clinical social workers using standardized evaluations and grouped into five domains: (1) economic hardship, (2) healthcare access barriers, (3) neighborhood/built environment instability, (4) social support challenge, and (5) Psycho-behavioral instability. We used multivariable adjusted Cox models to estimate hazard ratios (HRs) of all-cause, cardiovascular, and infection-related mortality; and logistic regression to estimate odds ratios (ORs) of 6-month rehospitalization risk.

**Results:** Among participants 1044 (62.9% males, mean age: 61.6 years), 601 (58%) reported at least 1social adversities: economic hardship (n=130), limited healthcare access (n=155), unstable housing (n=129), social support challenge (n=179), or psycho-behavioral instability (n=438). Over a mean follow up of 3.8 years, exposure to any SA was associated with higher all-cause mortality (HR 4.32; 95% confidence interval [CI] 3.03–6.14), CV mortality (HR 4.05; 95% CI 2.17–6.83), and infection-related mortality (HR 2.37; 95% CI 1.23–4.56). Social support challenge (HR 2.19; 95% CI 1.35–3.55) and psycho-behavioral instability (HR 1.96; 95% CI 1.24–3.11) were associated with higher CV mortality; economic hardship (HR 2.40; 95% CI 1.22–4.70) and social support challenge (HR 3.09; 95% CI 1.75–5.48) were associated with higher infection-related mortality. Compared with patients without SA, those with an environment instability, psycho-behavioral instability, or social support challenges had a 73% (aOR 1.73; 95% CI, 1.15–2.06), 75% (aOR 1.75; 95% CI, 1.31–2.35), and 44% (aOR 1.44; 95% CI, 1.00-2.06) higher risk of rehospitalization within 6 months, respectively.

**Conclusion:** SA was significantly associated with mortality and rehospitalization among PLHIV with HF, with domain-specific pathways influencing specific outcomes. A multidimensional assessment of social vulnerability may be useful to risk-stratify HF mortality risk in PLHIV.

## Introduction

People living with HIV (PLHIV) are increasingly surviving into older adulthood due to widespread use of antiretroviral therapy (ART), but this success has been accompanied by a growing burden of non-AIDS comorbidities, particularly cardiovascular diseases such as heart failure (HF)^1,2^. Heart failure now represents one of the most common and life-threatening chronic conditions among PLHIV, with significantly higher incidence, hospitalization rate, and mortality compared to HIV-negative individuals^3,4^. Prior research from the World Health Organization suggests that social determinants of health (SDoH) may explain up to 30% to 55% of chronic disease health outcomes by influencing lifestyle, behavior, stress, and environmental exposures^5^.

Barriers to optimal HF care are multifactorial but are especially pronounced in socioeconomically marginalized populations—a group disproportionately represented among PLHIV^6,7^. Social and behavioral risk factors such as housing instability, lack of insurance, food insecurity, and active substance use not only undermine medication adherence but also limit access to health services, including cardiology specialist care.^8^ Studies show that patients facing greater social adversity (SA) experience worse health outcomes^5,9^ and are less likely to receive lifesaving interventions like left ventricular assist devices^10,11^ or timely cardiology follow-up^8^. Emerging studies show that incorporating SDoH into predictive models improves mortality prediction in HF beyond clinical factors alone, suggesting the profound impact of these non-clinical variables^12^. Those findings emphasize the need for clinical frameworks that incorporate real-world SDoH assessment. Despite this, most studies to date lack validated, standardized SDoH assessment tools and struggle to capture the multidimensional nature of SA^9^. To our knowledge, none have longitudinally evaluated the impact of multidimensional social adversity on outcomes in PLHIV with HF.

To address this gap, we analyzed data from the NYC Health + Hospitals HIV–Heart Failure (NYC4H) cohort, a prospective, multicenter study of 1,044 PLHIV with HF across New York City’s public hospital system—the largest U.S. safety-net network. Our primary objective was to examine the impact of multidimensional SA, assessed via structured evaluations by licensed clinical social workers (LCSW), on all-cause, cardiovascular, and infection-related mortality. We also explored whether differences in SA exposure, care implementation and medication adherence mediated or modified these associations. By leveraging real-world data from a high-risk population, we aimed to identify actionable social targets for improving HF outcomes in PLHIV.

## Methods

### Study population

This study utilized data from the NYC 4H cohort, consolidating records from eleven major New York City Health + Hospitals.^13^ The dataset integrates retrospective baseline data collection with ongoing prospective follow-up. The original NYC 4H cohort comprised adult individuals with confirmed HIV and HF from inpatient or clinic visits between July 2017 and June 2022. HIV and HF diagnoses were identified via International Classification of Diseases, 10th Revision, Clinical Modification (ICD-10-CM) codes and confirmed by lab results and echocardiograms. Baseline demographics, medical history, treatments, and social factors were obtained through chart review at enrollment. The study was approved by the Institutional Review Board of NYC Health + Hospitals.

### Clinical variables

Self-reported baseline demographic information was retrieved from the electronic health record (EHR).^13^ Past medical histories were compiled through electronic chart reviews. Medication information for both baseline and follow-up were similarly obtained from clinical encounter records in the EHR, with additional cross-checking against internal or external pharmacy records to resolve discrepancies. Controlled HIV was defined as having an HIV viral load less than 200 copies/mL) at baseline. ART adherence was assessed by LCSW in the comprehensive evaluations. Guideline Directed Medical Therapy (GDMT) for HF is defined as prescription of renin–angiotensin system inhibitor, evidence-based beta-blocker, mineralocorticoid receptor antagonist (MRA) and sodium-glucose cotransporter 2 (SGLT2) Inhibitors. Functional status was assessed using Activities of Daily Living (ADL), with patients categorized as independent (0 deficits), partially dependent (1–2 deficits), or fully dependent (>2 deficits).

### Social adversities

SA were assessed through a psychosocial evaluation conducted during clinical care by LCSW during the initial clinic or hospital encounter within the enrollment period, as an integrated component of HIV-HF care. These comprehensive evaluations were conducted routinely to identify high risk patients and to address potential social needs. The comprehensive evaluation required approximately 45 minutes for a one-to-one personal interview. The spectrum of SA encompassed various factors, including criminal history, lack of insurance, immigration status, educational barriers, transportation barriers, financial or job instability, food insecurity, lack of family and community support, housing/living issues, active/past substance abuse, mental illness, home safety, history of trauma, identification with the Lesbian, Gay, Bisexual, Transgender and Queer (LGBTQ) community.

To enable interpretation and facilitate domain-level analysis, the 14 individual social and behavioral adversity variables were grouped into five conceptual categories based on a combination of SDoH frameworks from the Centers for Disease Control and Prevention (CDC) and other studies of SDoH in cardiovascular disease^14,15^: (1) Economic hardship (financial insecurity, job instability, food insecurity); (2) Healthcare access barriers (lack of insurance, immigration status, low educational attainment, transportation barriers); (3) Environment instability (housing instability, home safety concerns); (4) Social support challenges (lack of family/community support, LGBTQ+ identification); and (5) Psycho-behavioral instability (history of trauma, mental illness, polysubstance use, criminal history). Individuals could report multiple social adversities (SA) and thus be included in multiple domain-specific analyses.

### Outcomes

The Patient Outreach Department conducted annual follow-up attempts to schedule in-person visits for individuals who had missed routine HIV care. In this study, loss to follow-up was defined as the inability to reach a patient during the follow-up period despite these annual contact efforts. A total of 26 (2.5%) patients were lost to follow-up after a single encounter (details in cohort profile)^13^. Missing data were minimal and key clinical variables such as baseline CD4, viral load, EF had ≤3% missingness. For all adjusted covariates, complete case analysis was performed, and individuals with missing values were excluded from multivariable models.

Our primary outcome focused on the incidence of overall mortality during the follow-up period, initially identified through the EHR. Between November 2023 and December 2023, all patients or their family members were contacted to verify survival status. Patients were followed from baseline (date of first clinic or inpatient visit from July 2017 to June 2000) until death, loss to follow up, or censoring on October 30^th^ 2023.

Cardiovascular mortality was defined as death occurring during hospitalization with a primary medical problem listed as heart failure, myocardial infarction, ventricular arrhythmia, or cardiogenic shock. Causes of mortality were reviewed by two independent physicians. In case of disagreement between the primary reviewers, the cause of death was discussed by the entire research committee to achieve consensus. Additional secondary outcomes included infection related mortality through censor date and rehospitalization within 6 months of baseline date.

### Statistical analysis

Baseline characteristics were compared across grouped SA domains using descriptive statistics. Continuous variables were summarized as means ± standard deviation or medians with interquartile range, as appropriate, and categorical variables as counts with percentage. Between-group differences were assessed using one-way ANOVA or the Kruskal-Wallis test for continuous variables, and χ² tests for categorical variables.

All-cause mortality was compared by SA domains using Kaplan–Meier curves and log-rank tests. Multivariable Cox proportional hazards models were used to estimate hazard ratios (HRs) for mortality. Model 1 adjusted for age and sex; Model 2 further adjusted for baseline ejection fraction (EF), CD4 count, HIV control status, comorbidities (including hypertension, hyperlipidemia, diabetes, stroke, COPD, CKD, ESRD, cancer, HBV, HCV, and cirrhosis), heart failure etiology (ischemic vs non-ischemic), ADL, and smoking status. Proportional hazards assumptions were confirmed via Schoenfeld residuals without significant violations observed. Cardiovascular and infection related mortality outcomes were assessed in a similar manner, with Fine–Gray models being further implemented to account for competing risks.

Logistic regression models were used to estimate odds of 6-month rehospitalization. Model 1 adjusted for age and sex, while Model 2 included the full set of clinical covariates listed above. We tested the sensitivity of our all-cause mortality findings by restricting the analysis to the following clinically relevant subgroups: “HIV controlled patients”, “patients with reduced EF”, and “excluding events occurring in the first 3 months after baseline”. Effect modification was assessed by fitting an interaction term between the main SA domains as well as SA domains and the following potential modifiers (controlled HIV status, HF GDMT prescription at baseline, medication adherence, high co-morbidity burden). Mediation analyses examined whether GDMT at baseline, overall medication adherence, HIV medication adherence, or HIV control influenced the SA–mortality link in all domains.^16^ The confidence intervals for the indirect effects were estimated through bootstrap resampling in 500 iterations.

For the primary analyses and all other analyses, including mediation and moderation analyses, a p-value of <0.05 was considered as statistically significant. All analyses were conducted using Stata (version 15.1; StataCorp), R (version 4.3.3) and Python (version 3.1).

## Results

### Baseline Characteristics by Exposure to Social Adversity

Among the 1,044 participants (62.9% males, mean age: 61.6 years), 443 reported no social adversities, while 130 reported economic hardship, 155 experienced healthcare access barriers, 129 faced unstable living environments, 179 reported social support challenges, and 438 experienced psychosocial or behavioral instability. Over a mean follow-up period of 3.8 years, a total of 259 deaths were recorded, representing an overall mortality rate of 24.8%.

As shown in **Table 1**, key baseline differences emerged across groups. Higher proportions of male participants were observed in the domains of unstable environment (72.9%) and social support challenges (74.3%). Patients living in unstable environments and those suffering from psycho-behavioral instability were more likely to be active smokers (54.3% and 50.7%, respectively) compared with those without SA (29.1%). COPD prevalence followed a similar pattern. Baseline ASCVD prevalence was higher in patients who suffered from unstable living environments (63.6%) and economic hardship (62.3%), compared to those without SA (48.8%).

**Table 1.** Baseline characteristics of People Living With HIV and Heart Failure by social adversity domain.

|  | No SA | Economic hardship | Healthcare access barriers | Environment instability | Social support challenge | Psycho-behavioral instability |
| --- | --- | --- | --- | --- | --- | --- |
| **Participant Number** | 443 | 130 | 155 | 129 | 179 | 438 |
| **Average Age** | 58.8 | 58.3 | 59.8 | 59.1 | 60 | 59.8 |
| **Male, (%)** | 265 (59.8%) | 82 (63.1%) | 95% (61.3%) | 94 (72.9%) | 133 (74.3%) | 277 (62.9%) |
| **Race/Ethnicity** |  |  |  |  |  |  |
| Black/African American | 259 (58.5%) | 75 (57.7%) | 93 (60%) | 76 (58.9%) | 102 (57%) | 282 (64.4%) |
| Hispanic/Latino | 128 (28.9%) | 44 (33.9%) | 49 (31.6%) | 37 (28.7%) | 47 (26.2%) | 109 (24.9%) |
| Non-Hispanic white | 26 (5.9%) | 3 (2.3%) | 4 (2.6%) | 10 (7.6%) | 17(9.5%) | 22 (5%) |
| Asian & Pacific Islander | 13 (2.9%) | 1 (0.8%) | 3 (1.9%) | 1 (0.8%) | 2 (1.1%) | 3 (0.7%) |
| Other/Unknown | 17 (3.8%) | 7 (5.4%) | 6 (3.9%) | 5 (2.9%) | 11 (6.2%) | 22 (5%) |
| **Smoking** |  |  |  |  |  |  |
| Never smoker | 151 (34.1%) | 28 (21.5%) | 47 (30.3%) | 22 (17%) | 35 (19.6%) | 65 (14.6%) |
| Past smoker | 159 (35.9%) | 45 (34.6%) | 53 (34.2%) | 35 (27.1%) | 65 (36.3%) | 146 (33.3%) |
| Active smoker | 129 (29.1%) | 57 (43.9%) | 52 (33.6%) | 70 (54.3%) | 77 (43%) | 222 (50.7%) |
| **ADLs** |  |  |  |  |  |  |
| Completely independent | 320 (73.1%) | 92 (70.8%) | 113 (72.9%) | 91 (71.1%) | 114 (64.4%) | 301 (69.8%) |
| Partially dependent | 98 (22.4%) | 32 (24.6%) | 34 (21.9%) | 29 (22.7%) | 49 (27.7%) | 103 (23.9%) |
| Completely dependent | 20 (4.6%) | 6 (4.6%) | 8 (5.2%) | 8 (6.3%) | 14 (7.9%) | 27 (6.3%) |
| **Medical Comorbidities** |  |  |  |  |  |  |
| Hypertension | 343 (77.4%) | 92 (70.8%) | 114 (73.5%) | 96 (74.4%) | 124 (69.3%) | 323 (73.7%) |
| Type II diabetes mellites | 188 (42.4%) | 47 (36.2%) | 62 (40.3%) | 49 (38.3%) | 61 (34.3%) | 163 (37.3%) |
| Hyperlipidemia | 170 (38.4%) | 44 (38.9%) | 59 (38.1%) | 52 (40.3%) | 61 (34.1%) | 141 (32.2%) |
| ASCVD | 216 (40.5%) | 81 (62.3%) | 88 (56.8%) | 82 (63.6%) | 99 (55.3%) | 231 (52.7%) |
| Obesity (BMI >30) | 127 (28.7%) | 44 (33.8%) | 38 (24.5%) | 31 (24%) | 51 (28.5%) | 131 (29.9%) |
| Chronic kidney disease | 112 (25.3%) | 29 (22.3%) | 33 (21.3%) | 30 (23.3%) | 32 (17.9%) | 79 (18%) |
| End-stage renal disease | 54 (12.2%) | 25 (19.2%) | 30 (19.4%) | 17 (13.2%) | 31 (17.3%) | 48 (11%) |
| COPD | 158 (35.7%) | 40 (30.8%) | 48 (31%) | 56 (43.3%) | 71 (39.7%) | 195 (44.5%) |
| **HIV** |  |  |  |  |  |  |
| Prescribed ART at baseline | 412 (93%) | 108 (83.9%) | 134 (86.5%) | 112 (86.8%) | 156 (87.2%) | 390 (89%) |
| Non-compliance to ART | 43 (9.7%) | 47 (36.2%) | 38 (24.5%) | 56 (43.3%) | 45 (25.1%) | 98 (22.4%) |
| HIV viral control (<200c/ml) | 305 (68.8%) | 90 (69.2%) | 103 (66.5%) | 93 (72.1%) | 130 (72.6%) | 317 (72.4%) |
| Median CD4 count at baseline (cells/mm<sup>3</sup>) | 439 | 328 | 364 | 306 | 287 | 331 |
| **Heart failure** |  |  |  |  |  |  |
| Ischemic heart disease | 171 (38.6%) | 48 (36.9%) | 56 (36.1%) | 49 (38%) | 64 (35.8%) | 160 (36.5%) |
| Median EF at baseline (%) | 45 | 46 | 45 | 45 | 45 | 45 |
| Baseline >1 GDMT med | 302 (68.2%) | 69 (53.1%) | 96 (61.9%) | 63 (48.8%) | 91 (50.8%) | 234 (53.4%) |
| Baseline >2 GDMT med | 157 (35.4%) | 31 (23.9%) | 43 (27.7%) | 24 (18.6%) | 36 (20.1%) | 112 (25.6%) |
| Ischemic evaluation | 138 (31.2%) | 36 (27.7%) | 38 (24.5%) | 43 (33.3%) | 58 (32.4%) | 123 (28.1%) |
| Revascularization | 44 (9.9%) | 9 (6.9%) | 15 (9.7%) | 10 (7.8%) | 15 (8.4%) | 26 (5.9%) |

SA: Social adversities; ADL: Activities of daily living; ASCVD: Atherosclerotic cardiovascular disease; COPD: Chronic obstructive pulmonary disease; HIV: Human immunodeficiency virus; ART: Antiretroviral therapy; EF: Ejection fraction; GDMT: Guideline-directed medical therapy

Poor adherence to ART was significantly more common among participants experiencing SA and remained consistently elevated across all five domains 22.4-43% vs. 9.7% among those without any SA). Despite this, median CD4 counts and HIV viral loads were comparable, and rates of controlled HIV (HIV viral load < 200 copies/mL) were similar across all groups (69–76%, P=0.16). Ischemic evaluation was uncommon in general, but even less so for patients with healthcare access barriers (24.5%) compared to those without any SA (31.2%). Revascularization procedures were lowest in patients with psycho-behavioral instability (5.9%). Being prescribed at least one class of GDMT was less common in patients with environment instability (48.8%), social support challenges (50.8%) and psycho-behavioral instability (53.4%), compared to those without SA (68.2%).

Figure 1 illustrates the distribution of domain-level intersections among social adversities. The most common single-domain exposure was psycho-behavioral instability (n = 230, 22%), followed by healthcare access barriers (n = 62, 5.9%) and social support challenges (n = 44, 4.2%). The most frequent intersection of adversities occurred between social support challenges and psycho-behavioral instability (n = 56, 5.4%), followed by economic hardship combined with psycho-behavioral instability (n = 31, 3.0%). Notably, six of the ten most frequent individual or intersecting adversity exposures involved the psycho-behavioral instability domain.

**Figure 1.**
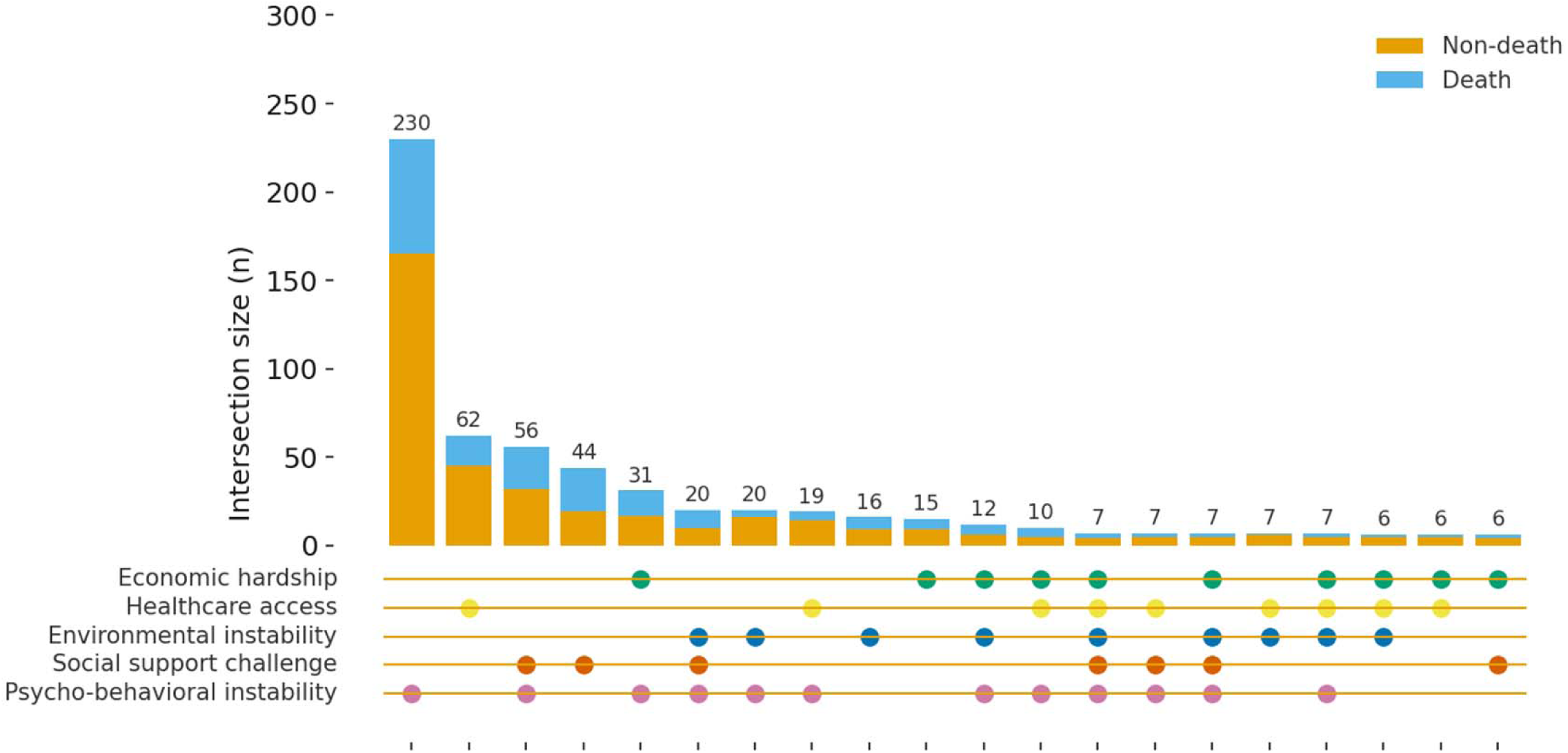
Figure 1 Distribution of deaths across intersecting domains of social adversity.

### Survival Outcomes and Rehospitalization by Social Adversity Domain

Survival curves for all domains diverged early and continued to separate over time [Figure 2], with log-rank tests confirming significant differences between SA exposure groups (P<0.05). After adjusting for covariates, having any social adversity was associated with a 4.32-fold (95% CI 3.03-6.16, P<0.001) increase in overall mortality [**Table 2**]. Compared with no SA, the presence of any SA was associated with a 3.92-fold higher risk of CV mortality (95% CI, 2.18-7.02, P<0.001) and a 2.68-fold higher risk of infection-related mortality (95% CI, 1.37-5.26, P<0.01). Among individual domains, social support challenges (HR 2.28; 95% CI, 1.41-3.68, P<0.01) and psycho-behavioral instability (HR 1.84; 95% CI, 1.15-2.92, P=0.01) were associated with higher CV mortality, while economic hardship (HR 2.48; 95% CI 1.26-4.9, P<0.01) and social support challenges (HR 3.19, 95% CI 1.79-5.68, P<0.001) were associated with higher infection-related mortality.

**Figure 2.**
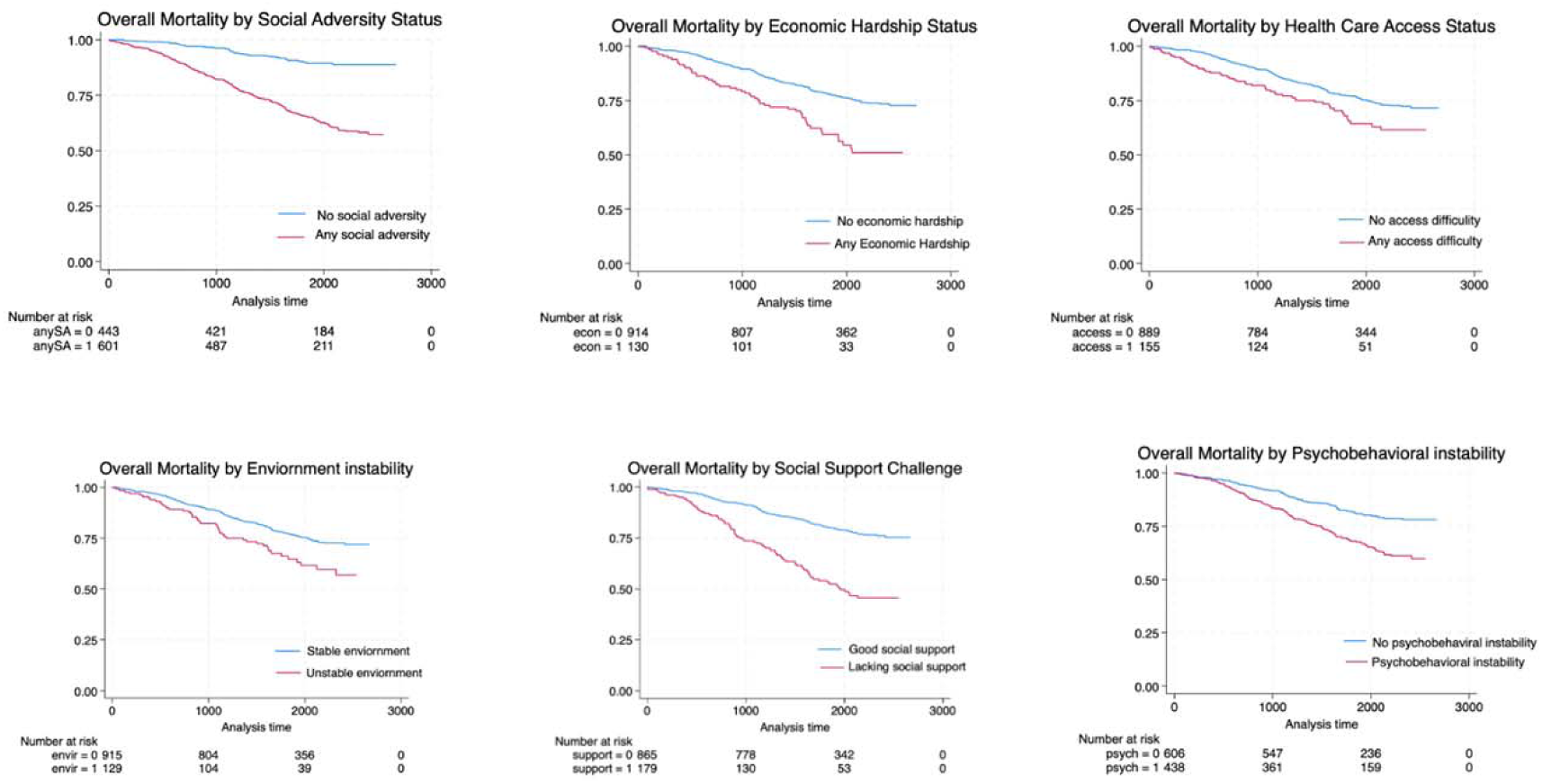
overall mortality in different Social Adversities domain.

**Table 2.**
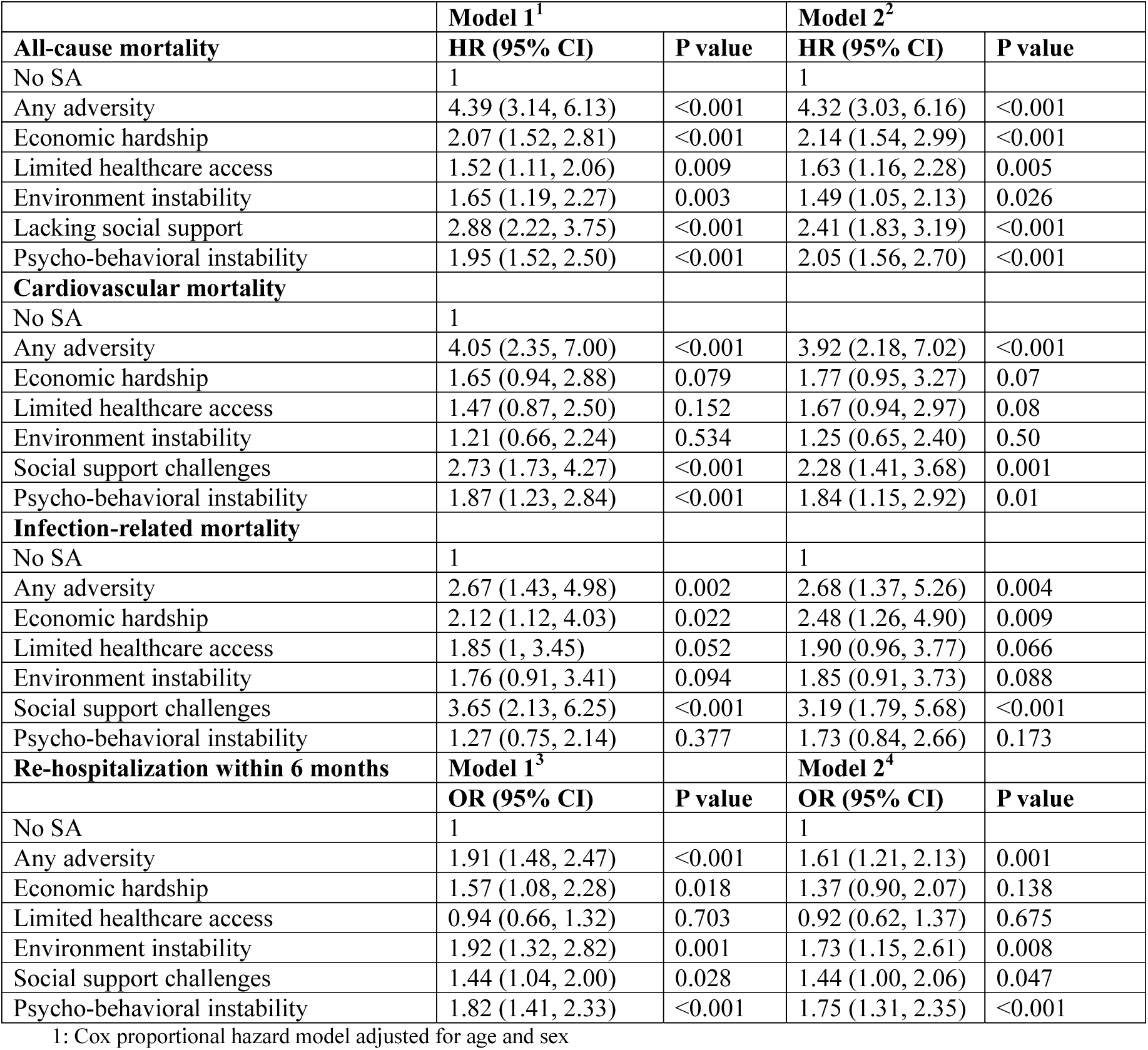
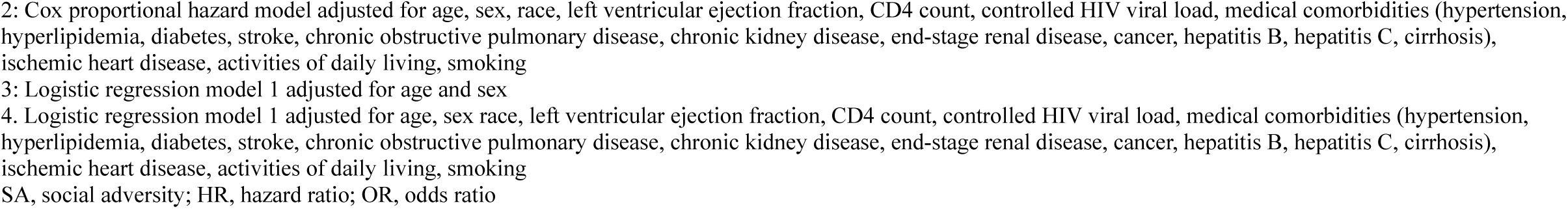
Mortality and rehospitalization outcomes by social adversity domain.

|  | **Model 1<sup>1</sup>** |  | **Model 2<sup>2</sup>** |  |
| --- | --- | --- | --- | --- |
| **All-cause mortality** | **HR (95% CI)** | **P value** | **HR (95% CI)** | **P value** |
| No SA | 1 |  | 1 |  |
| Any adversity | 4.39 (3.14, 6.13) | <0.001 | 4.32 (3.03, 6.16) | <0.001 |
| Economic hardship | 2.07 (1.52, 2.81) | <0.001 | 2.14 (1.54, 2.99) | <0.001 |
| Limited healthcare access | 1.52 (1.11, 2.06) | 0.009 | 1.63 (1.16, 2.28) | 0.005 |
| Environment instability | 1.65 (1.19, 2.27) | 0.003 | 1.49 (1.05, 2.13) | 0.026 |
| Lacking social support | 2.88 (2.22, 3.75) | <0.001 | 2.41 (1.83, 3.19) | <0.001 |
| Psycho-behavioral instability | 1.95 (1.52, 2.50) | <0.001 | 2.05 (1.56, 2.70) | <0.001 |
| **Cardiovascular mortality** |  |  |  |  |
| No SA | 1 |  |  |  |
| Any adversity | 4.05 (2.35, 7.00) | <0.001 | 3.92 (2.18, 7.02) | <0.001 |
| Economic hardship | 1.65 (0.94, 2.88) | 0.079 | 1.77 (0.95, 3.27) | 0.07 |
| Limited healthcare access | 1.47 (0.87, 2.50) | 0.152 | 1.67 (0.94, 2.97) | 0.08 |
| Environment instability | 1.21 (0.66, 2.24) | 0.534 | 1.25 (0.65, 2.40) | 0.50 |
| Social support challenges | 2.73 (1.73, 4.27) | <0.001 | 2.28 (1.41, 3.68) | 0.001 |
| Psycho-behavioral instability | 1.87 (1.23, 2.84) | <0.001 | 1.84 (1.15, 2.92) | 0.01 |
| **Infection-related mortality** |  |  |  |  |
| No SA | 1 |  | 1 |  |
| Any adversity | 2.67 (1.43, 4.98) | 0.002 | 2.68 (1.37, 5.26) | 0.004 |
| Economic hardship | 2.12 (1.12, 4.03) | 0.022 | 2.48 (1.26, 4.90) | 0.009 |
| Limited healthcare access | 1.85 (1, 3.45) | 0.052 | 1.90 (0.96, 3.77) | 0.066 |
| Environment instability | 1.76 (0.91, 3.41) | 0.094 | 1.85 (0.91, 3.73) | 0.088 |
| Social support challenges | 3.65 (2.13, 6.25) | <0.001 | 3.19 (1.79, 5.68) | <0.001 |
| Psycho-behavioral instability | 1.27 (0.75, 2.14) | 0.377 | 1.73 (0.84, 2.66) | 0.173 |
| **Re-hospitalization within 6 months** | **Model 1<sup>3</sup>** |  | **Model 2<sup>4</sup>** |  |
|  | **OR (95% CI)** | **P value** | **OR (95% CI)** | **P value** |
| No SA | 1 |  | 1 |  |
| Any adversity | 1.91 (1.48, 2.47) | <0.001 | 1.61 (1.21, 2.13) | 0.001 |
| Economic hardship | 1.57 (1.08, 2.28) | 0.018 | 1.37 (0.90, 2.07) | 0.138 |
| Limited healthcare access | 0.94 (0.66, 1.32) | 0.703 | 0.92 (0.62, 1.37) | 0.675 |
| Environment instability | 1.92 (1.32, 2.82) | 0.001 | 1.73 (1.15, 2.61) | 0.008 |
| Social support challenges | 1.44 (1.04, 2.00) | 0.028 | 1.44 (1.00, 2.06) | 0.047 |
| Psycho-behavioral instability | 1.82 (1.41, 2.33) | <0.001 | 1.75 (1.31, 2.35) | <0.001 |

1: Cox proportional hazard model adjusted for age and sex
2: Cox proportional hazard model adjusted for age, sex, race, left ventricular ejection fraction, CD4 count, controlled HIV viral load, medical comorbidities (hypertension, hyperlipidemia, diabetes, stroke, chronic obstructive pulmonary disease, chronic kidney disease, end-stage renal disease, cancer, hepatitis B, hepatitis C, cirrhosis), ischemic heart disease, activities of daily living, smoking
3: Logistic regression model 1 adjusted for age and sex
4. Logistic regression model 1 adjusted for age, sex race, left ventricular ejection fraction, CD4 count, controlled HIV viral load, medical comorbidities (hypertension, hyperlipidemia, diabetes, stroke, chronic obstructive pulmonary disease, chronic kidney disease, end-stage renal disease, cancer, hepatitis B, hepatitis C, cirrhosis), ischemic heart disease, activities of daily living, smoking
SA, social adversity; HR, hazard ratio; OR, odds ratio

Multivariable logistic regression revealed that exposure to any SA was associated with a 61% higher risk of re-hospitalization within 6 months (adjusted odds ratio [aOR] 1.61, 95% CI: 1.21–2.13, *P* = 0.001) [**Table 2**]. Compared with patients without SA, those with an environment instability, psycho-behavioral instability, or social support challenges had a 73% (aOR 1.73; 95% CI, 1.15–2.06), 75% (aOR 1.75; 95% CI, 1.31–2.35), and 44% (aOR 1.44; 95% CI, 1.00-2.06) higher risk of rehospitalization within 6 months, respectively.

Effect-modification analyses suggested significant interactions between economic hardship and environmental instability (p-value = 0.006), economic hardship and social support challenge (P=0.012), as well as social support challenges and psycho-behavioral instability (p <0.001). Furthermore, the pairwise correlation between the individual SA domains is shown in **Supplement** Figure 1 and **Supplement** Figure 2. Significant effect modification was noticed between psych-behavioral instability and high burden of co-morbidity. Mediation analysis showed that being prescribed ≥2 GDMT drugs at baseline had a significant indirect effect (95% CI 0.006-0.146), showing a potential pathway to overall mortality. **(Supplement Table 3)**

### Specific adversities and risk of mortality/ re-hospitalization

Figure 3 demonstrates the association between individual domains of SA and risk of combined mortality or re-hospitalization. Having social support challenges (HR 2.94, 95% CI: 2.25–3.85, *P* < 0.01), food insecurity (HR 2.87, 95% CI: 1.73–4.76, *P* < 0.01), employment instability (HR 1.92, 95% CI: 1.25–2.95, *P* < 0.01), active polysubstance use (HR 1.89, 95% CI: 1.48–2.42, *P* < 0.01), transportation barriers (HR 1.79, 95% CI: 1.20–2.67, *P* < 0.01), financial instability (HR 1.72, 95% CI: 1.14–2.60, *P* = 0.01), housing insecurity (HR 1.65, 95% CI: 1.18–2.42, *P* < 0.01), and a history of mental illness (HR 1.59, 95% CI: 1.22–2.08, *P* < 0.01) were all independently associated with higher all-cause mortality. In terms of re-hospitalization within 6 months, significant associations were observed in patients with a history of trauma (OR 2.23, 95% CI: 1.21–4.09, *P* = 0.01), active polysubstance use (OR 1.75, 95% CI: 1.32–1.94, *P* < 0.01), and housing insecurity (OR 1.65, 95% CI: 1.11–2.44, *P* < 0.01).

**Figure 3.**
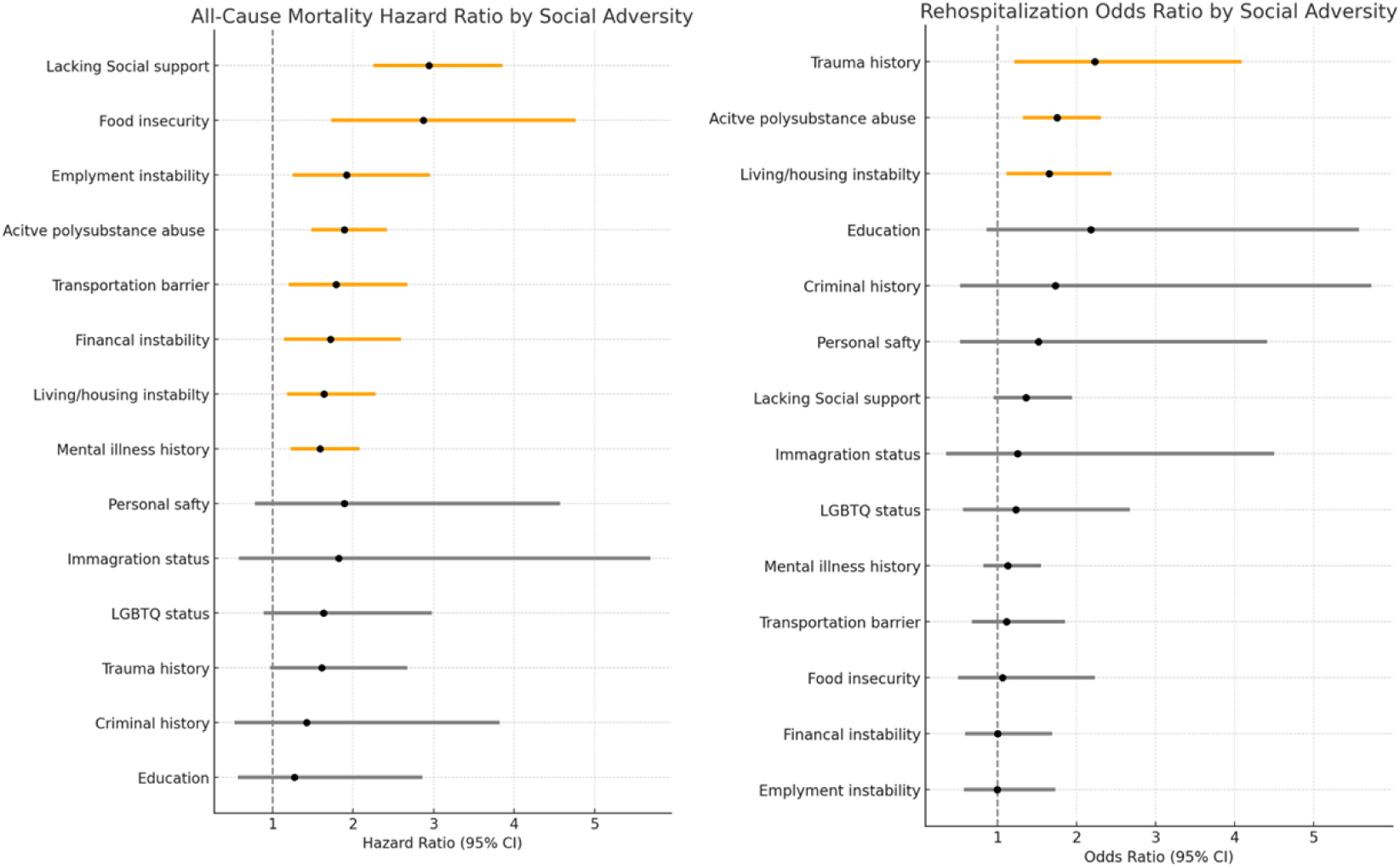
Forest plot for mortality and re-hospitalization across individual social adversities.

## Sensitivity Analyses

Sensitivity analyses confirmed the robustness of primary findings across clinically relevant subgroups. Lag analysis using a 90-day threshold showed similar associations. Fine-Gray competing risk models for cardiovascular and infection-related mortality yielded comparable results (**Supplementary Table 4-5**).

## Discussion

To our knowledge, this is the largest real-world cohort of PLHIV and HF in which multi-dimensional SA was systematically evaluated by licensed clinical social workers using standardized assessments. This approach provides a rare, comprehensive view into the lived realities of patients navigating chronic disease amid overlapping structural, psychosocial, and behavioral challenges. In this cohort, the presence of social adversity was associated with significantly higher risks of all-cause, cardiovascular, and infection-related mortality, as well as hospital readmission. Notably, individual SA domains demonstrated distinct patterns of association with specific outcomes, suggesting that different types of adversity may act through unique and potentially targetable pathways.

While prior research in the field has demonstrated that social determinants such as housing instability, food insecurity, and social support challenge are independently associated with adverse cardiovascular outcomes^7,17–19^, very few have multi-dimensional data to fully evaluate their effect in the context of multimorbidity. Notably, the association between social adversity and mortality in our cohort was stronger than that reported in studies of either HIV or heart failure alone, suggesting that the impact of social adversity may be amplified in individuals living with both conditions.^20,21^ The chronicity and self-management demands of coexisting HIV and HF—each requiring high levels of care coordination, medication adherence, and system navigation—may render this population especially sensitive to social adversity^22–24^. Taken together, these results emphasize that the interplay of biomedical complexity and social adversity is not merely additive but synergistic—amplifying vulnerability in ways underappreciated by traditional risk models focused on single disease frameworks.

In our study, distinct domains of social adversity demonstrated specific associations with clinical outcomes, underscoring the heterogeneity of risk pathways^24,25,26,27^. Social support challenges and psycho-behavioral instability were linked to markedly higher cardiovascular mortality, suggesting that disruptions in social connectedness and behavioral stability may exacerbate the complexity of heart failure management^24,25^. In contrast, economic hardship and social support challenges were most strongly associated with infection-related mortality, highlighting the role of material deprivation and isolation in limiting timely access to HIV care, infection prevention and treatment. Furthermore, environment instability and psycho-behavioral instability predicted substantially higher 6-month rehospitalization rates, reflecting the destabilizing effect of environmental and behavioral factors on post-discharge management^26,27^.

Although conventional domain-specific analyses of social determinants of health (SDoH) demonstrate that different social adversities contribute to poor outcomes through distinct pathways, frontline providers often encounter patients with multiple, overlapping adversities that do not fit neatly into a single category. Traditional binary or checklist-style assessments evaluate each SA domain independently, which may obscure the real-world complexity of co-occurring challenges. Our findings underscore the importance of standardized, multidimensional SA assessments to evaluate the real social burden and guide targeted interventions. Without structured tools, informal or inconsistent documentation risks systematically underestimating patient vulnerability, limiting opportunities for early intervention and risk-guided care.

Our mediation analyses provide additional insights into the mechanisms by which SA leads to worse clinical outcomes. Notably, ART adherence was consistently lower across all five domains among individuals with SA, in contrast to those without SA. In our study, this reduced adherence did not lead to lower rates of HIV viral suppression, perhaps highlighting the robustness of the modern ART regimens or perhaps a liberal definition of viral control (<200c/ml). We believe inconsistent ART use may still contribute to low level viremia, inflammation and subsequently worse clinical outcomes. In parallel, rates of GDMT use may partially mediate the relationship between SA and adverse outcomes, indicating that proper initial treatment could mitigate the adverse effect of SA on clinical outcomes. These findings highlight important opportunities for targeted interventions, particularly enhancing medication adherence and ensuring early, appropriate GDMT to improve outcomes in this vulnerable population.

The present study demonstrates how SA impact clinical outcomes in a vulnerable and biologically complex population living with HIV and HF. Our findings also highlight the importance of detailed multi-domain assessment of SA in this high-risk population and opportunities for targeted interventions based on specific SA domains to improve outcomes. Future research should refine practical approaches for assessing SA in routine HIV clinical care to enable more precise risk stratification and tailored interventions for PLHIV with HF and other chronic co-morbidities. Wrap around services to address SA are already a focus of many large academic HIV clinics in the USA. These services include robust social work support and case management, food assistance, co-located substance use and mental health treatment, among others. Such services are often supported with supplemental Ryan White funding from the US Health Services Research Administration.^28^ As a result, HIV clinics may be better equipped to address social determinants of health than similar primary care clinics for people without HIV. Opportunities exist to leverage these resources to specifically address HF care for PLHIV with a high burden of SA.

## Limitation

This study has several limitations. First, its observational design limits causal inference. Although we found strong associations between SA and mortality in extensively adjusted regression models, unmeasured confounders may partially explain the results. Second, SA was only assessed at baseline, so we could not account for dynamic changes in social circumstances over time. A third limitation is the possibility of reverse causation. Severe illness could lead to SA. Despite these concerns, the consistency in lag analysis and dose-response nature of the association supports a likely causal contribution of social adversity to mortality.

## Conclusion

Domains and individual SAs were strongly associated with increased mortality and re-hospitalization among with PLHIV and HF, with domain-specific pathways influencing specific outcomes. A multidimensional assessment of social vulnerability could be useful to risk-stratify HF mortality risk in PLHIV. Our findings should inform the development of future biobehavioral interventions and implementation research to improve outcomes for PLHIV and HF.

## Author contribution statement

YC conceived the research question, study design, data analysis, and manuscript preparation. PB, NN and MP contributed to data collection. LB contributed to data analysis and manuscript preparation. YC, PB, NN, LB, MP, CV, IC, JB, VG, SB all contributed to data collection. RF and LP supervised data collection and manuscript preparation. ESB, SE and CTL provided additional interpretation of study findings and supervised manuscript preparation.

The guarantor of the study is Yi-Yun Chen; accepts full responsibility for the finished work and/or the conduct of the study, had access to the data, and controlled the decision to publish.

## Data Availability

Data described in the manuscript, code book, and analytic code will be made available upon request pending application and approval.

## Acknowledgements

The authors wish to extend their sincere gratitude to the staff and participants of the NYC 4H cohort for their invaluable contributions.

## Ethical Compliance

This study was deemed to be exempt from the institutional review board at the Albert Einstein College of Medicine. All data were collected retrospectively and discarded upon completion of the study.

---

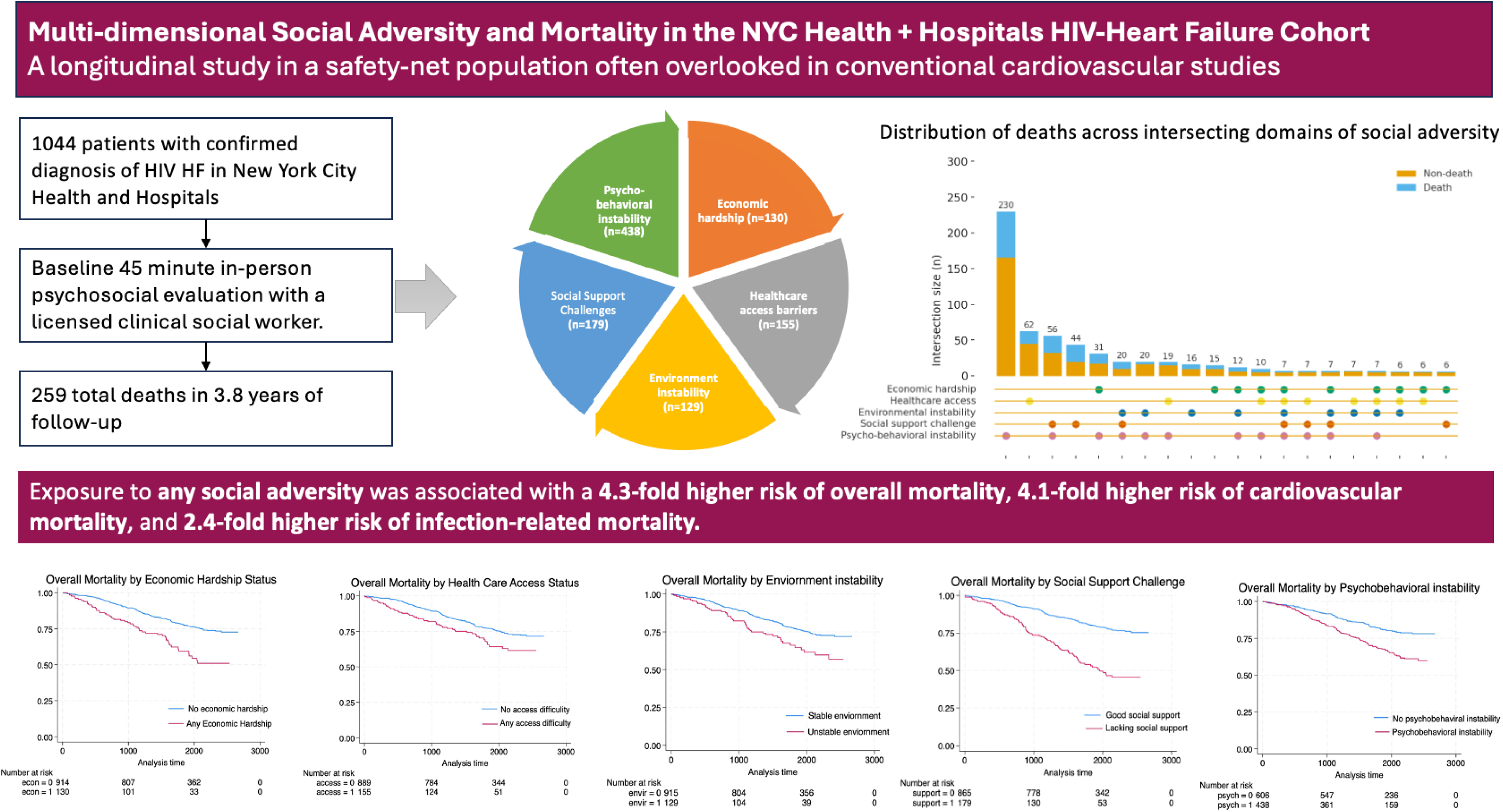

